# Endotoxemia and circulating bacteriome in severe COVID-19 patients

**DOI:** 10.1101/2020.05.29.20109785

**Authors:** Phatadon Sirivongrangson, Win Kulvichit, Sunchai Payungporn, Trairak Pisitkun, Ariya Chindamporn, Sadudee Peerapornratana, Prapaporn Pisitkun, Suwalak Chitcharoen, Vorthon Sawaswong, Navaporn Worasilchai, Sarinya Kampunya, Opass Putcharoen, Thammasak Thawitsri, Nophol Leelayuwatanakul, Napplika Kongpolprom, Vorakamol Phoophiboon, Thitiwat Sriprasart, Rujipat Samransamruajkit, Somkanya Tungsanga, Kanitha Tiankanon, Nuttha Lumlertgul, Asada Leelahavanichkul, Tueboon Sriphojanart, Terapong Tantawichien, Usa Thisyakorn, Chintana Chirathaworn, Kearkiat Praditpornsilpa, Kriang Tungsanga, Somchai Eiam-Ong, Visith Sitprija, John A. Kellum, Nattachai Srisawat

**Author notes:** Equal contribution for this work (Phatadon Sirivongrangson, Win Kulvichit). Corresponding author: Nattachai Srisawat, MD, PhD Division of Nephrology, Department of Medicine, Faculty of Medicine, King Chulalongkorn Memorial Hospital, Bangkok 10330, Thailand.

## Abstract

**Purpose:** When severe, COVID-19 shares many clinical features with bacterial sepsis. Yet, secondary bacterial infection is uncommon. However, as epithelium are injured and barrier function is lost, bacterial products entering the circulation might contribute to the pathophysiology of COVID-19.

**Methods:** We studied 19 adults, severely ill patients with COVID-19 infection, who were admitted to King Chulalongkorn Memorial Hospital, Bangkok, Thailand, between 13^th^ March and 17^th^ April 2020. Blood samples on day 1, 3, and 7 of enrollment were analyzed for endotoxin activity assay (EAA), (1→3)-β-D-Glucan (BG), and 16S rRNA gene sequencing to determine the circulating bacteriome.

**Results:** Of the 19 patients, 14 were in intensive care and 10 patients received mechanical ventilation. We found 8 patients with high EAA (≥ 0.6) and about half of the patients had high serum BG levels which tended to be higher in later in the illness. Although only 1 patient had a positive blood culture, 18 of 19 patients were positive for 16S rRNA gene amplification. Proteobacteria was the most abundant phylum. The diversity of bacterial genera was decreased overtime.

**Conclusions:** Bacterial DNA and toxins were discovered in virtual all severely ill COVID-19 pneumonia patients. This raises a previously unrecognized concern for significant contribution of bacterial products in the pathogenesis of this disease

## INTRODUCTION

While most cases coronavirus disease 2019 (COVID-19) are mild, severe COVID-19 pneumonia can occur with a mortality rate as high as 50% [1]. It is unclear why some patients develop clinical features of sepsis/septic shock with multiple organ dysfunction [2]. The majority of bacterial cultures from severe COVID-19 patients are negative, [3] and although empiric antibiotics are commonly used, [3-5] they are not recommended [6]. However, while the respiratory tract is the principle site of infection for COVID-19, the disease has been shown to involve the GI tract as well and symptoms such as diarrhea are reported in about a third of cases [7]. Enterocytes in ileum and colon express the ACE2 receptor and virus has been detected in stool. Thus, there is a possibility that bacterial translocation from the GI tract might complicate severe COVID-19 disease [8].

Endotoxin, a part of the cell wall of Gram-negative bacteria, has been extensively investigated and acknowledged as one of the key triggers of lethal shock during severe sepsis and also one of the primary drivers of the cytokine storm [9-11]. Serum (1 ➔ 3)-b-D-glucan (BG) has been evaluated as a potential marker of intestinal barrier dysfunction. Serum BG was tested in several mouse models of gut leakage, including dextran sulfate solution administration, endotoxin injection, and cecal ligation and puncture sepsis [12]. However, the presence of endotoxemia and serum BG in severe COVID-19 have never been examined. Not only bacterial toxin but also direct bacterial invasion might play role in severe COVID-19. Exploring circulating bacteriome in severe COVID-19 may allow us to test the presence of any bacterial invasion during critical illness.

Thus, we designed this investigation to determine whether bacterial products were present in the blood of severe COVID-19 pneumonia patients and whether their source was likely to be the gut as evidenced by serum BG. We also sought to characterize the circulating bacteriome in COVID-19 pneumonia.

## METHODS

### Study population, setting, and data collection

This was a prospective observational study in COVID-19 pneumonia patients admitted to King Chulalongkorn Memorial Hospital, Bangkok, Thailand, between 13^th^ March and 17^th^ April 2020. Our inclusion criteria included (1) age >18 years, (2) confirmed COVID-19 pneumonia, and (3) had leftover blood samples. The first day of enrollment was the day that patients fulfilled inclusion criteria. The study was reviewed and approved by Faculty of Medicine, Chulalongkorn University ethics committee (IRB no. 336/63). The informed consent was waived due to the observational nature of the study. The study was designed and conducted according to the STROBE guideline [13].

We obtained demographic data, information on clinical presentations, laboratory investigations, and radiography at the time of presentation, and during intensive care unit (ICU) admission. We collected blood samples that were left over on day 1, day 3, and day 7 after enrollment. All laboratory tests and radiologic assessments, including standard chest radiographs and chest computed tomography, were performed at the discretion of the treating physician. Endotoxin activity assay (EAA), cytokines, and serum BG were measured on day 1, 3, and 7 of enrollment. We assessed clinical outcomes on day 28 after enrollment, including mechanical ventilation, ventilator-free day, vasopressor, prone position, extracorporeal membrane oxygenation (ECMO), acute kidney injury (AKI), renal replacement therapy (RRT), and successful extubation.

### Study definitions

A confirmed case of COVID-19 was defined by a positive result of a reverse-transcriptase-polymerase-chain-reaction (RT-PCR) assay of a specimen collected from a nasopharyngeal swab. We defined COVID-19 pneumonia as a COVID-19 case who showed the evidence of pulmonary infiltration from chest radiography or chest computer tomography. We defined severe COVID-19 as a COVID-19 case who was admitted in ICU. Acute kidney injury was defined based on serum creatinine and urine output criteria according to the Kidney Disease Improving Global Outcome 2012 guideline [14]. We defined patient with high endotoxin by EAA ≥0.6 on day 1.

### Standard of care treatment

Treating physicians performed thorough evaluations and managed COVID-19 patients with standard care including volume status assessment, hemodynamic and respiratory support according to Surviving Sepsis Campaign: Guidelines on the Management of Critically Ill Adults with Coronavirus Disease 2019 (COVID-19) [6]. Although, currently, there is no specific antiviral treatment for COVID-19, antiviral therapy was given to all patients with confirmed COVID-19 pneumonia as recommended by the Department of Medical Service, Ministry of Public Health of Thailand. The treatment consists of a combination of (1) favipiravir, (2) lopinavir/ritonavir or darunavir, (3) hydroxychloroquine, and (4) azithromycin.

### Laboratory procedures

#### COVID-19 test confirmation

COVID-19 tests were performed by qRT-PCR technique using cobas^®^ SARS-CoV-2 qualitative test with the cobas^®^ 6800 platform (Roche Diagnostics, Indianapolis, IN). We followed the manufacturer’s instructions for testing. The samples were obtained by nasopharyngeal swab and preserved in viral transport cases before sending to analysis. The qRT-PCR tests provided cycle threshold (Ct) value for each test. These values represent the number of cycles required for the positive fluorescent signal. Therefore, the lower Ct values correlated with the higher viral load. The assay is designed to detect ORF1 genes and N genes of SARS-CoV-2. The result was considered positive when the Ct values of both target genes were < 40, negative when they were both >40. If only one of the target genes had a Ct value < 40, the tests were confirmed by another RT-PCR machine, CFX96 Touch qPCR Detection System (Bio-Rad, Hercules, CA) with detection of 3 genes (ORF1 gene, ORF3 gene, and N gene). All procedures were performed in a biosafety level 2 laboratory.

#### Endotoxin activity assay

We performed the chemiluminescent-based endotoxin activity assay (EAA; Spectral Diagnostics, Ontario, Canada) as described elsewhere [15]. This assay is based on the detection of enhanced respiratory burst activity in neutrophils following their priming by complexes of endotoxin and a specific anti-endotoxin antibody. Briefly, 40 μL of whole blood were incubated with zymosan and anti-endotoxin antibody. The endotoxin activity level of ≥0.60 was considered as high activity level.

#### Serum (1 ➔ 3)-b-D-glucan (BG)

We analyzed serum for BG with Fungitell^®^ assay (Associates of Cape Cod, Falmouth, MA) following manufacturer instructions. Briefly, 5 mL of serum was mixed with an alkaline pretreatment reagent (0.125 M KOK/0.6 M KCl) and incubated at 37°C for 10 minutes. One hundred microliters of the reconstituted Fungitell reagent was added to each well and the reaction monitored at A405nm–A490nm for 40 minutes. The Fungitell assay detects BG through the activation of factor G, a protease zymogen which activates a second protease zymogen, pro-clotting enzyme, that cleaves a chromophore from a chromogenic peptide resulting in light absorbance at 405 nm. Serum BG >60 pg/mL was used as positive cut-off [16]. BG values at <7.8 pg/mL and >523.4 pg/mL were recorded as 0 and 523 pg/mL, respectively.

#### Cytokines

We measured a panel of cytokines including Interleukin 1 beta (IL-1β), Interferon alpha-2 (IFN-α2), Interferon gamma (IFN-γ), Tumor necrosis factor-α (TNF-α), Monocyte chemoattractant protein-1 (MCP-1), Interleukin 6 (IL-6), Interleukin 8 (IL-8), Interleukin 10 (IL-10), Interleukin-12, p70 (IL-12 p70), Interleukin17A (IL-17A, Interleukin 18 (IL-18), Interleukin 23 (IL-23), and Interleukin 33 (IL-33) at the same time points using LEGENDplex™ Human Th Cytokine Panel (BioLegend, San Diego, CA, USA) according to manufacturer’s protocol.

#### 16S rDNA high-throughput sequencing

Genomic DNA was extracted from 300 µl of whole blood using the GenUp_TM_ gDNA kit (Biotechrabbit, Germany). The amplification of the bacterial 16S rDNA was performed in total volume 25 µl consisting of *Taq* DNA Polymerase (0.5U) (Biotechrabbit, Germany), 1.5 mM MgCl_2_, 0.2 mM dNTPs, 0.2 mM forward primer 5’-ACTCCTACGGRAGGCAGCAG-3’ and 0.2 mM reverse primer 5’CCGTCAATTYYTTTRAGTTT-3’. The PCR product was re-amplified within V4 region of 16S rDNA by using phasing adaptor primers following from [17]. Amplified PCR products (~430bp) were purified by using the QIAquick PCR Purification Kit (QIAGEN, Germany) and quantified by KAPA library quantification kits for Illumina platforms (Kapa Biosystems, USA). The DNA libraries were pooled at equal amount and paired-end (2×250 cycles) sequenced on an Illumina MiSeq platform with a MiSeq Reagent Kit V2 (Illumina, USA).

### Statistical analyses

Raw sequencing data were demultiplexed by MiSeq reporter software (version 2.6.2.3). The FASTQ files were analyzed by QIIME2 pipeline (version 2019.7) [18]. The paired-end sequences were merged and trimmed based on quality score (<Q30). Then, merged reads were deduplicated and clustered with 99% similarity by using VSEARCH [19]. UCHIME algorithm were used for filtered out the chimeric sequences [20]. The filtered reads were classified based on 16S Greengene database [21] using VSEARCH algorithm. The alpha diversity was analyzed by implemented function in QIIME2. Differential abundance analysis was conducted by Linear discriminant analysis Effect Size (LEfSe) [22]. Wilcoxon matched pairs test were analyzed using GraphPad Prism version 6.01.

Statistical comparisons for continuous and categorical data were performed using Chi-square/Fisher Exact test and Mann-Whitney U test/Kruskal-Wallis test. Data is reported as counts (percentages) for categorical and median with interquartile range for continuous data. No imputation was performed on missing data. All statistical analyses were performed using Stata version 15.1(STATA Corp, TX). P value of less than 0.05 was considered as statistical significance for all tests performed.

## RESULTS

A total of 147 patients were recruited. Of these, 53 (34.6%) patients were diagnosed COVID-19 pneumonia. Among patients with COVID-19 pneumonia, only 19 patients fulfilled the inclusion criteria and 13 (68.4%) patients were admitted in the ICU (Figure 1). Male sex was predominant in our cohort. Markers of inflammations including ferritin, C-reactive protein (CRP), and IL-6 were markedly high (Table 1). We showed detailed clinical data and outcomes of 19 patients in Table 2 and Table S1.

**Figure 1.**
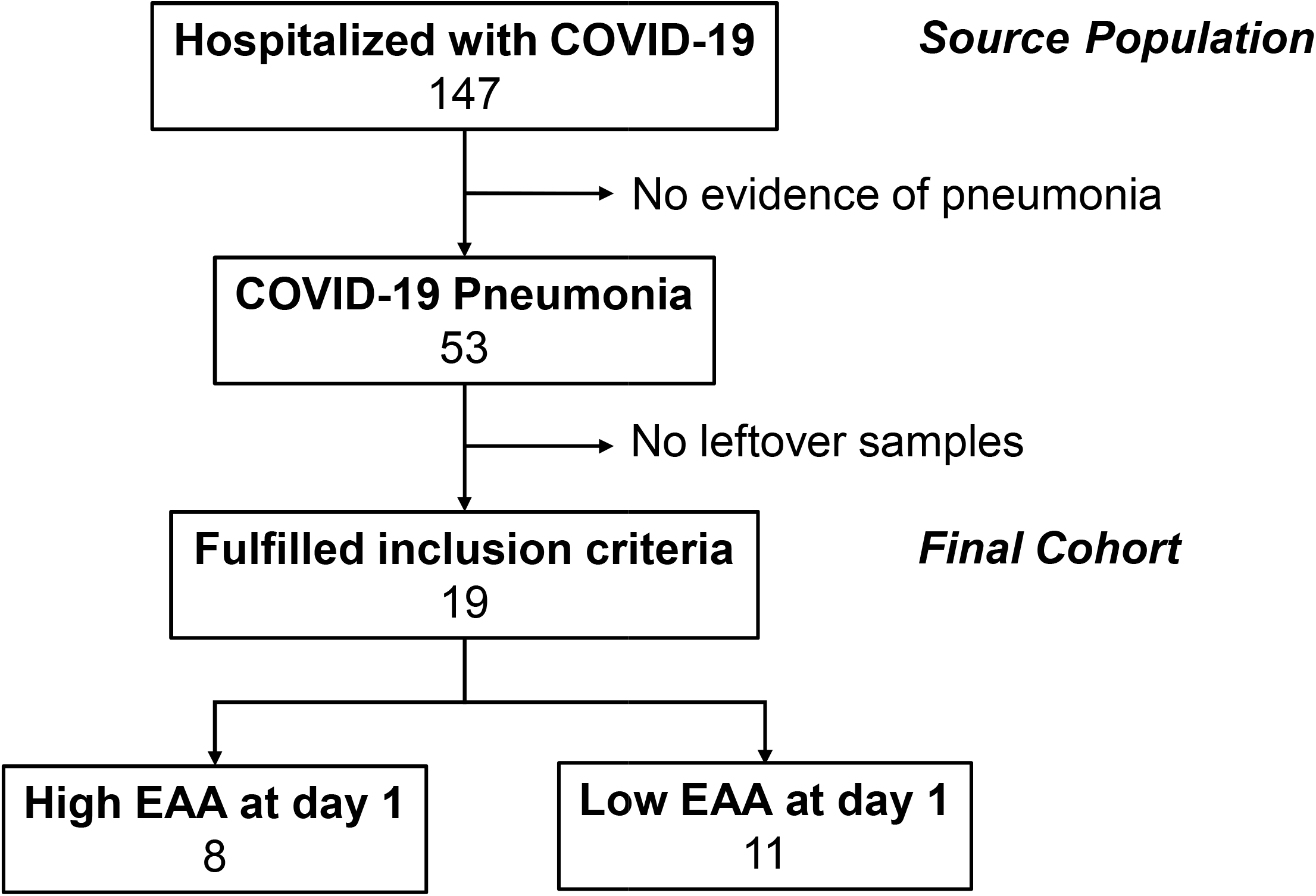
Study cohort.

**Table 1.**
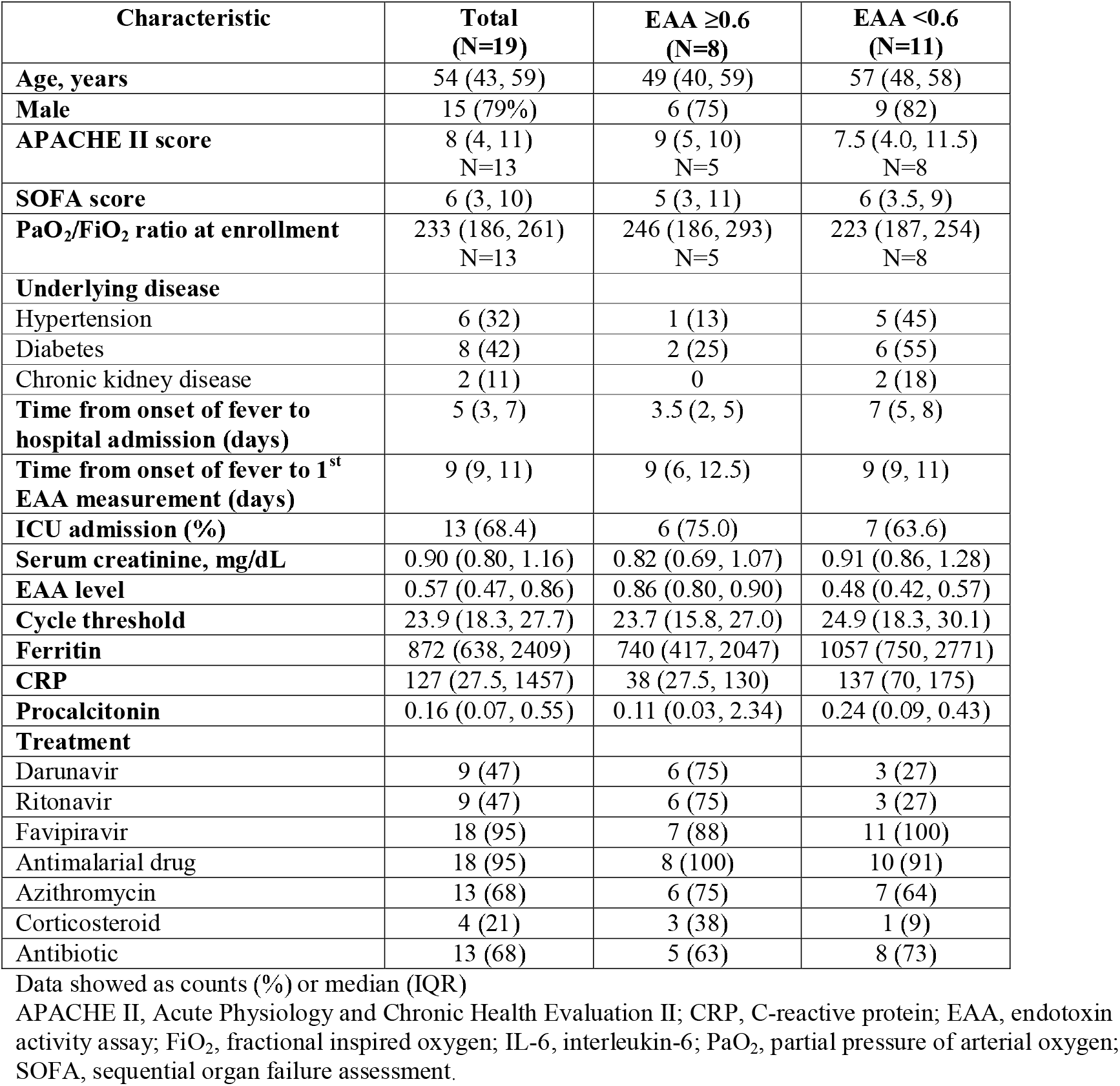
Clinical characteristics of COVID-19 pneumonia at enrollment

### Endotoxin activity and BG

Overall median (IQR) EAA on day 1, day 3, and day 7 were 0.57 (0.47, 0.86), 0.65 (0.49, 0.96), and 0.57 (0.43, 0.74) respectively. There were 8 (42.1%) patients with day 1 EAA ≥ 0.60 (Table 1). Distributions of EAA on day 1, day 3, and day 7 are shown in Figure 2a.

**Figure 2a.**
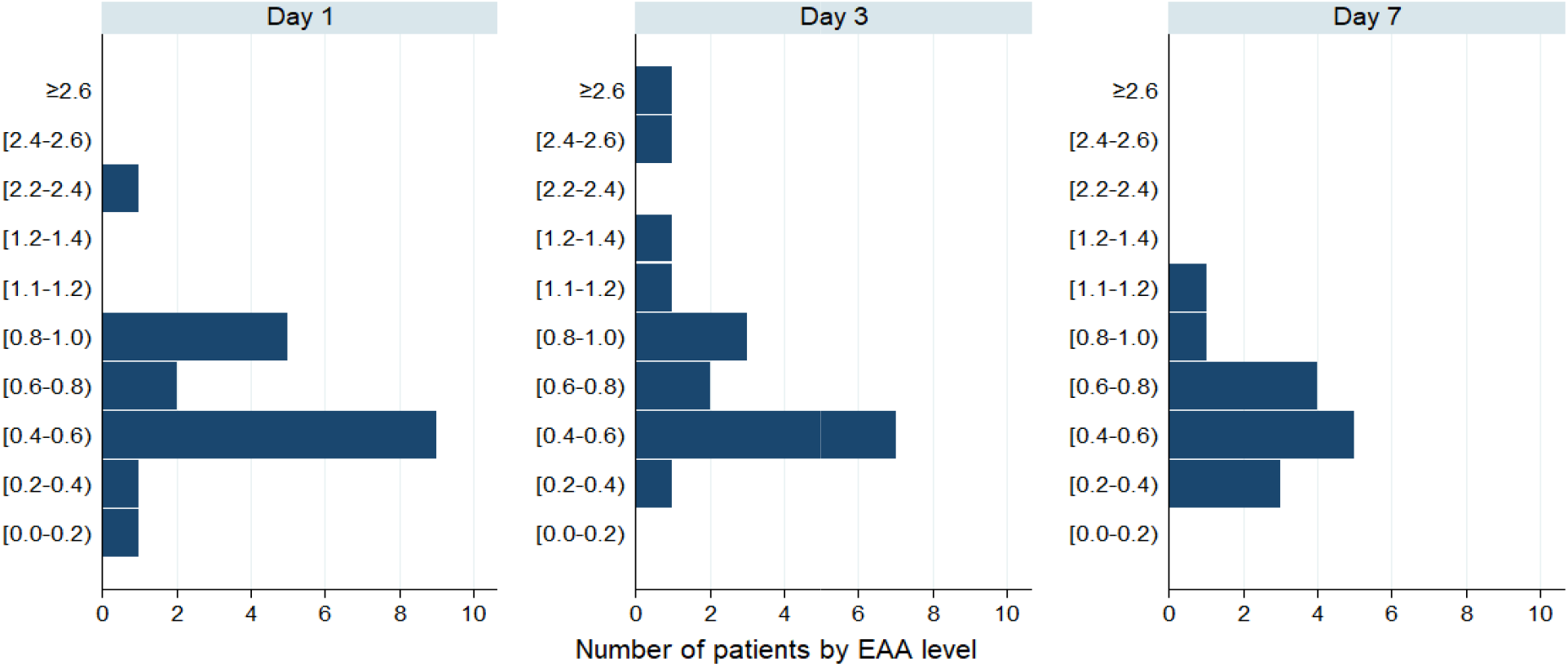
EAA distribution of COVID-19 pneumonia.

Figure 2b demonstrates the distributions of serum BG following day of enrollment. During 7 days of observation, 8 patients (42.1%) had high BG levels (defined as BG >60 pg/mL), and 37.5% of patients with high BG also had EAA >0.6. The level of BG increased following the day of enrollment (Figure 2b). The median BG levels were higher in patients with high EAA compared those with low EAA [day1 (25 VS12 pg/mL), day 3 (42 vs 10 pg/mL), and day 7 (75 vs 26 pg/mL)].

**Figure 2b.**
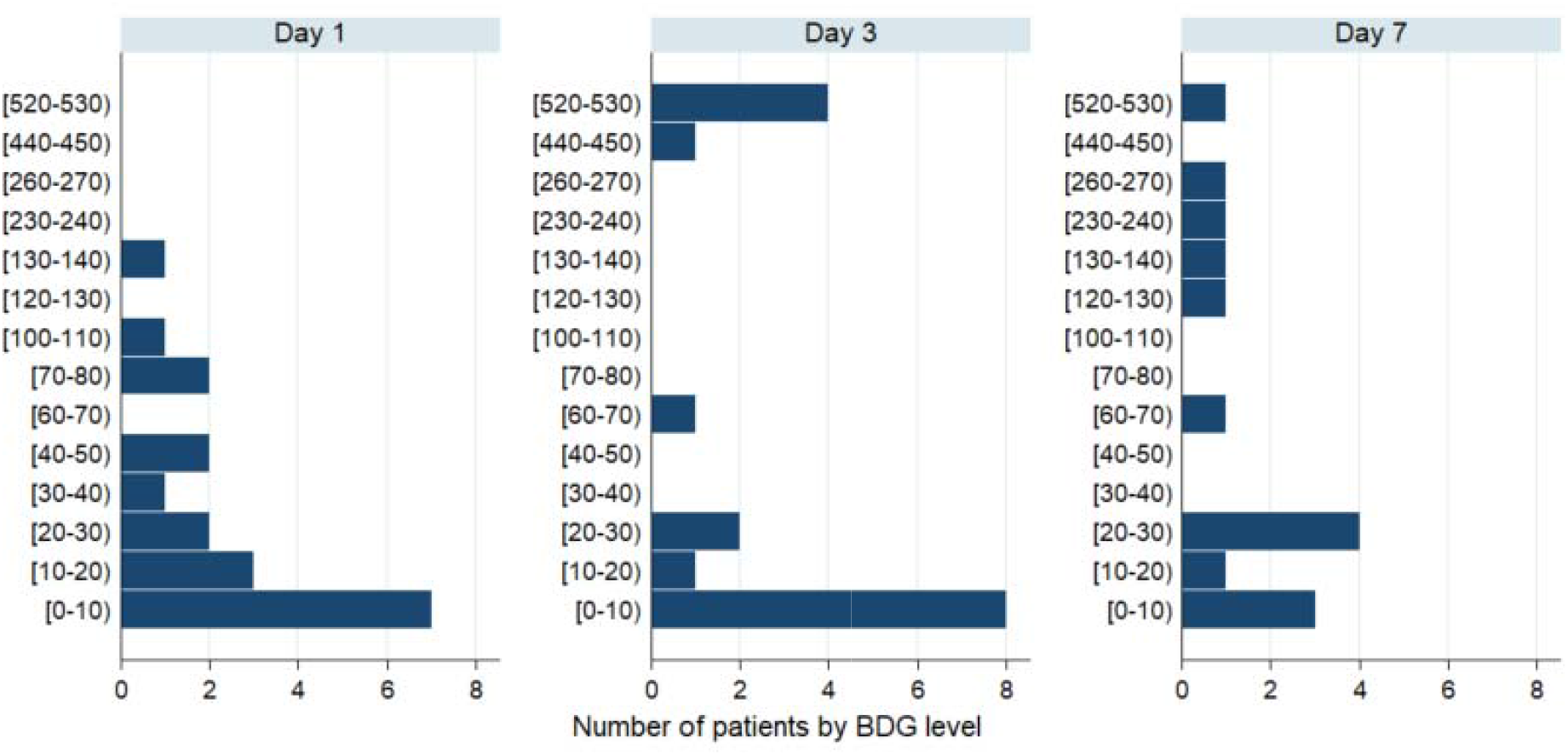
Serum BG distribution of COVID-19 pneumonia.

### Circulating bacteriome

Forty-nine of 50 samples (98.3%) from 19 COVID-19 pneumonia patients had presence of bacterial DNA in serum. Rarefaction curves were shown in Figure S1. Bacterial classification revealed that the relative abundances of bacteria were different among patients and days of illness (Figure 3). Taxonomy composition phyla abundances revealed that the Proteobacteria phylum was the predominant phylum at every time point (Figure 4, S2). Interestingly, Gram-negative bacterial genera including *Sphingomonas*, *Bradyrhizobium*, *Enhydrobacter*, *Phyllobacterium*, *Agrobacterium*, *Comamonas, Sediminibacterium*, *Acinetobacter, and Pseudomonas* were most likely found in different days of illness (Figure 3). Biodiversity of bacteria, demonstrated by the Chao1 richness, on day 1 was significantly higher than on day 3 (Figure S3, S4). The bacterial genera including *Sphingomonas* and *Sediminibacterium* were significantly (P<0.05) higher on day 3 compared to day 1; whereas *Comamonas*, *Acinetobacter*, and *Pseudomonas* were significantly (P<0.05) decreased on day 3 (Figure S5).

**Figure 3.**
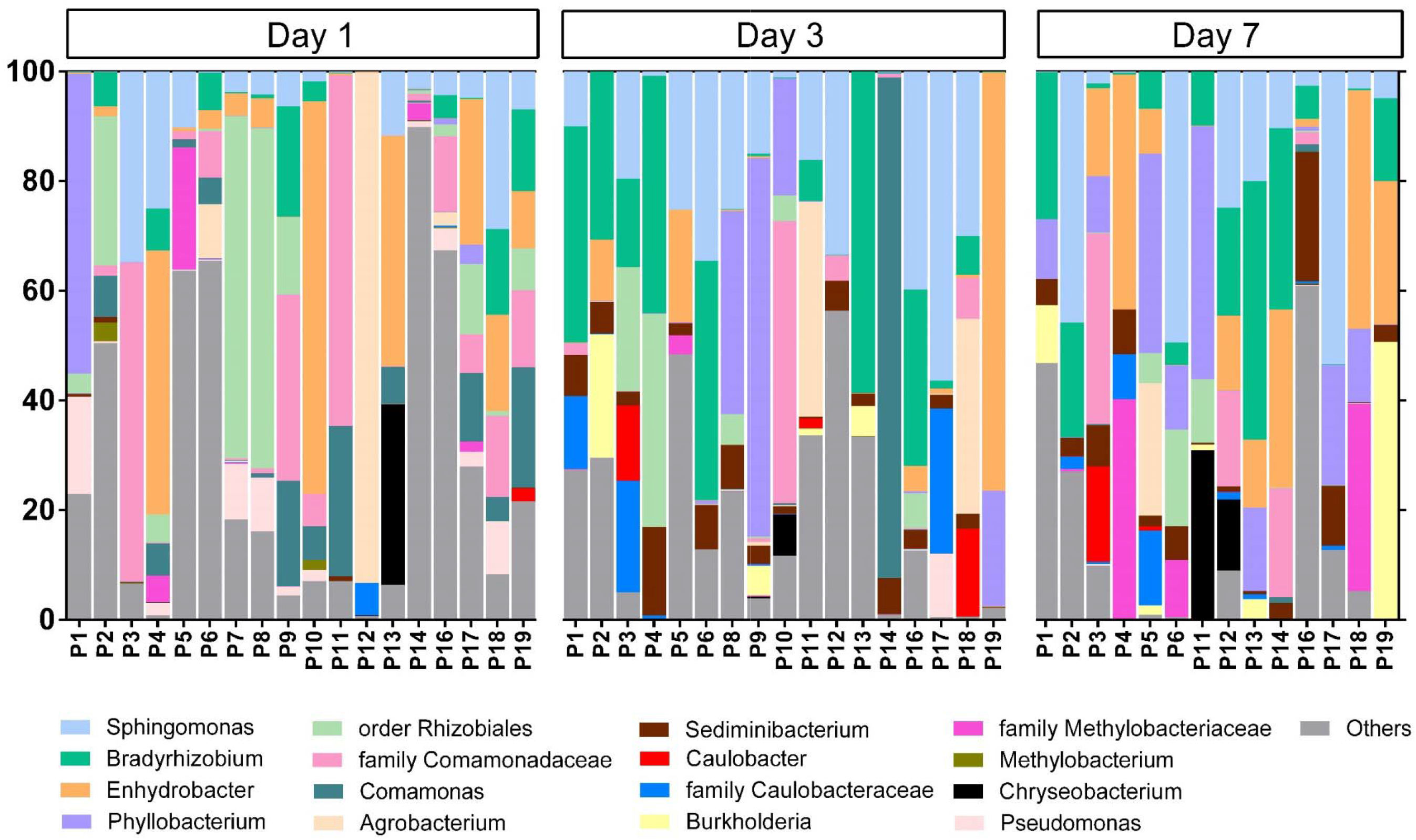
Dynamic bacterial community profiles on day 1, day 3, and day 7

**Figure 4.**
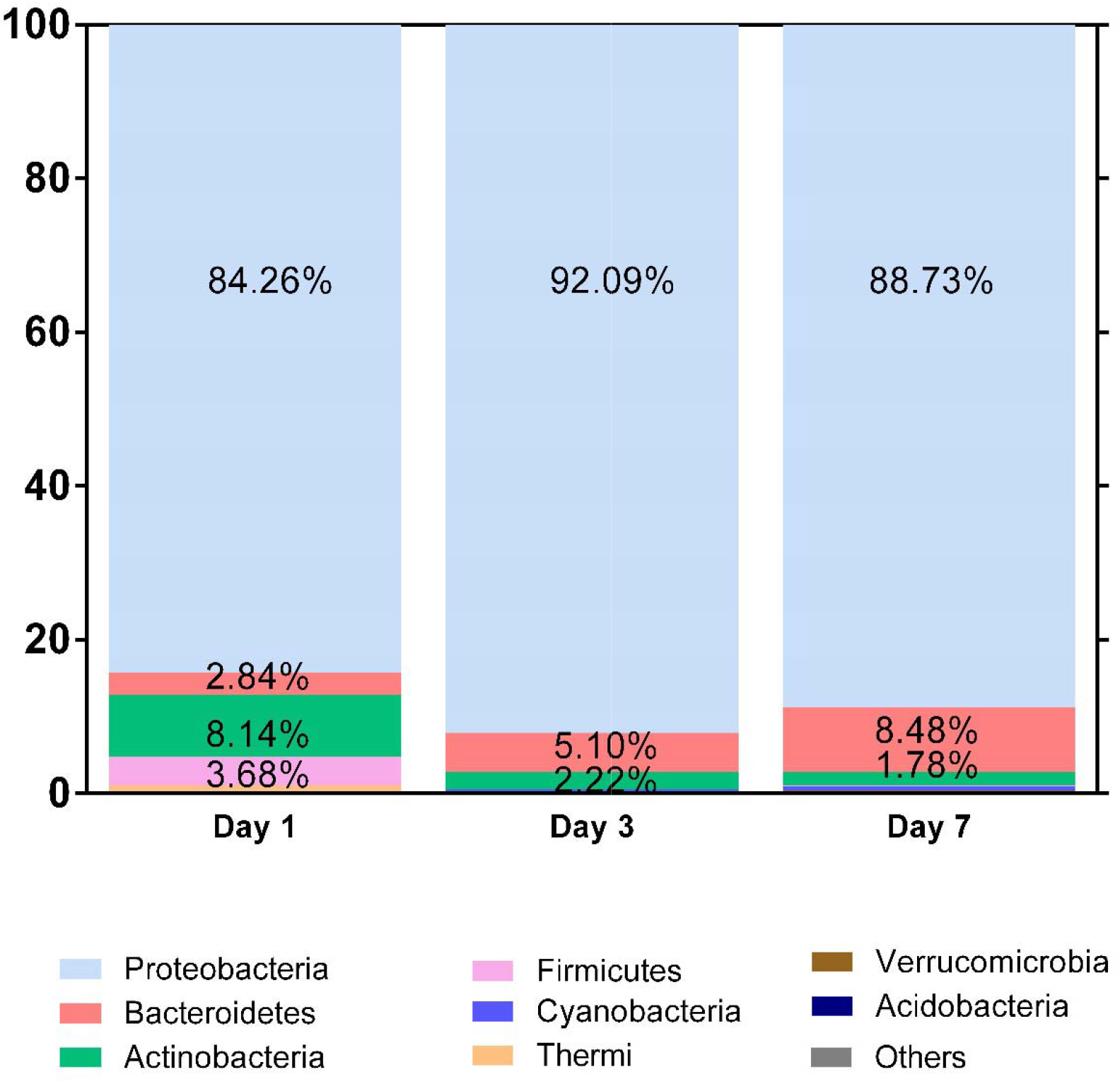
Overall summary of dynamic bacterial community profiles in COVID-19 patients on day 1, 3, and 7. Proteobacteria were observed to be the dominant bacterial phyla followed by Bacteroidetes, Actinobacteria and Firmicutes.

### Cytokines

A heat map of cytokine levels on day 1, 3, and 7 is depicted in Figure 5. Most of COVID-19 pneumonia had elevated of cytokines. On day 1, MCP-1, IL-6, IL-8, IL-10 in COVID-19 pneumonia who were admitted in ICU were significantly higher than COVID-19 pneumonia who were not admitted in ICU [(848 (410, 1782) vs 285 (215, 300), P=0.003; 45 (16, 334) vs 12 (7, 25), P=0.023; 73 (47, 128) vs 24 (20, 41), P=0.009; 23 (19, 56) vs 8 (2, 13), P=0.022.] (Table S1).

**Figure 5.**
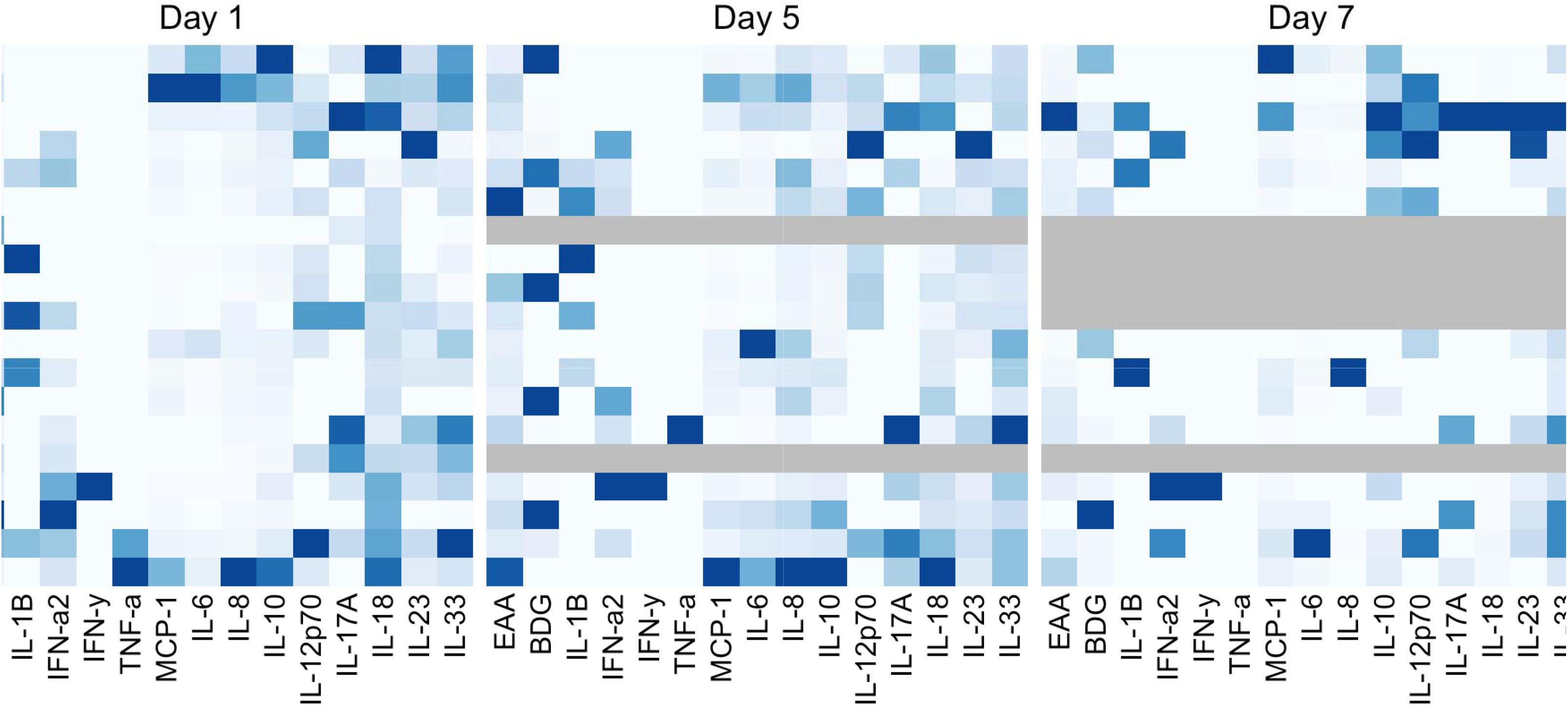
Cytokine heat map on day 1, day 3, and day 7

### Clinical characteristics at baseline and outcomes

Comparing clinical features between patients with high (≥0.6 on day 1) and low EAA (< 0.6 on day 1) revealed that those with high EAA had lower median age, and higher severity scores (Table 1). Patients with high EAA levels sought medical attention earlier than patients with low EAA levels [median time from onset of fever to hospital admission [3.5(2, 5) vs 7 (5, 8), P =0.01].

Regarding clinical outcomes, patients with high EAA were more likely to develop subsequent bacterial infection within 28 days after enrollment (38% vs 18%). Patients with high EAA tended to need more mechanical ventilation support than the low EAA group, although the statistical significance was not attained (62.5% vs 45.5%, P=0.65). The proportion of patients requiring vasopressors, prone position, and ECMO did not differ between EAA groups (Table 2).

**Table 2.**
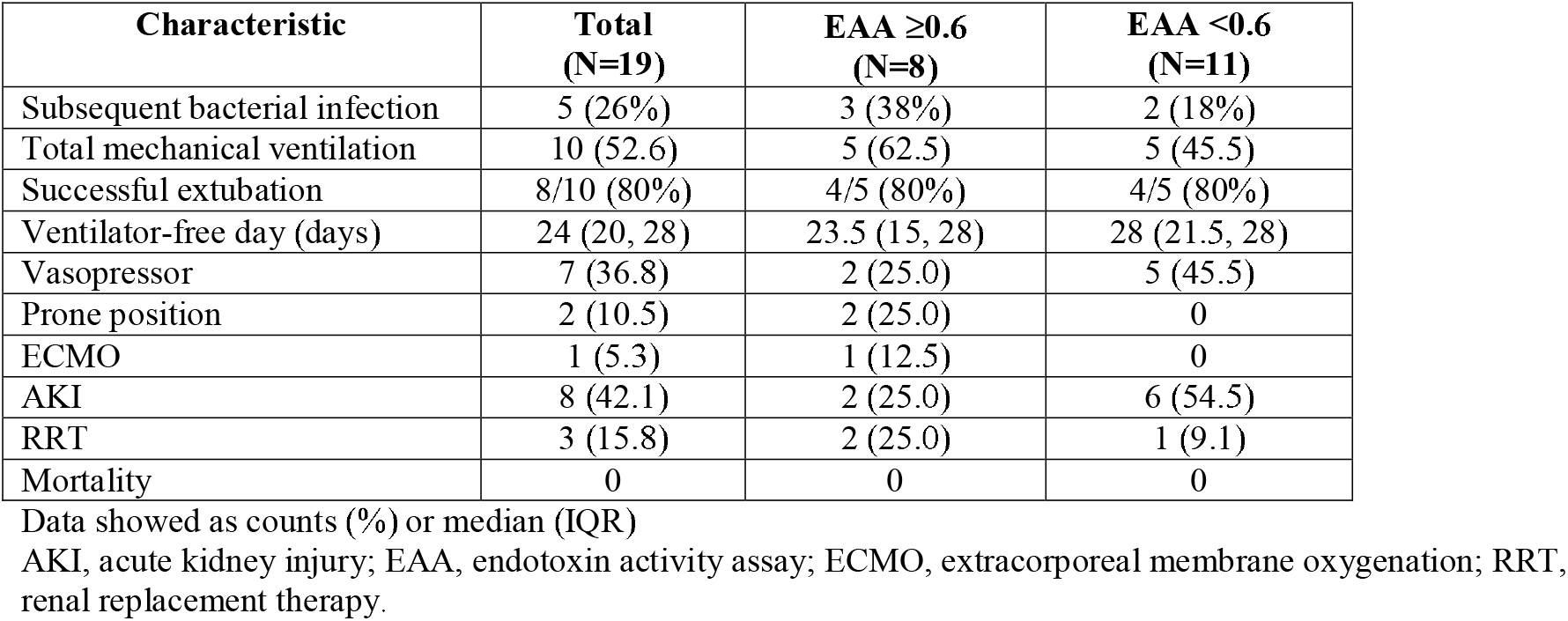
Outcomes at 28 days after enrollment

Twenty-eight-day outcomes are shown in Figure 6. The overall rate of AKI (any stage) in our study was 42.1%. Three (15.8%) patients required renal replacement therapy (RRT). When compared to patients with low EAA, patients with high EAA tended to have lower ventilator-free days [23.5 (15, 28) vs 28 (21.5, 28); P=0.51] and successful extubation rate (20% vs 60%; P=0.52). No patient died within 28 days after enrollment.

**Figure 6.**
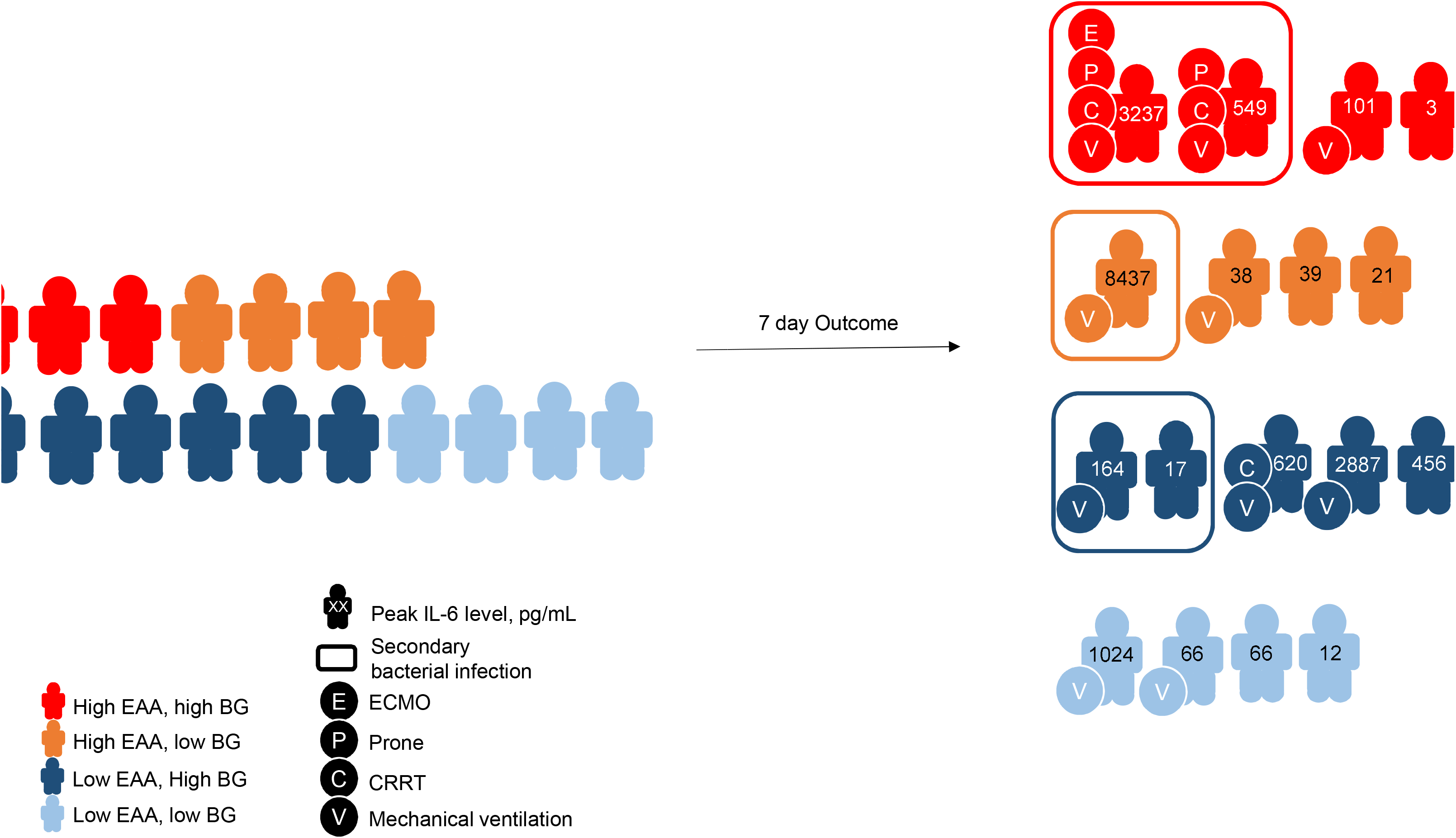
Clinical outcomes in study cohort.

## Discussion

In this cohort of COVID-19 pneumonia, we show that nearly 90% and 40% of patients had endotoxemia (defined as a moderate to high level of EAA), and high levels of BG, a measure of gut permeability (Figure 2). Using Next Generation Sequencing (NGS), we could also demonstrate the dominant bacterial DNA came from Proteobacteria, a phylum of Gram-negative bacteria which includes several pathogens that can cause sepsis, (Figure 4).

In animals, high endotoxin activity in viral infection has been demonstrated to be associated with poor outcome. Inoculation of Lipopolysaccharides (LPS) in mice with influenza infection resulted in the activation of local pulmonary inflammatory responses and may lead to secondary bacterial pneumonia [23]. LPS binds with toll-like receptor 4 (TLR4) and activates transcription factors activating protein-1 (AP-1), nuclear factor kappa B (NF-kB) and interferon regulatory factor 3 (IRF3) through myeloid differentiation factor 88 (MyD88) and TIR-domain-containing adapter-inducing interferon-β (TRIF)-dependent pathways. This leads to the induction of proinflammatory cytokines and interferons [24].

BG is a key structural polysaccharide of the cell wall of most fungi including *Candida spp*., and has been used in the diagnosis of invasive fungal infection [16, 25]. Leelahavanichkul et al. reported the use of serum BG as a biomarker of gut permeability in human sepsis [12]. In our study, the increase of serum BG level later in the course of illness (Figure 2b) might reflect overgrowth of *Candida spp*. in the gastrointestinal tract from antibiotics exposure [26]. Intestinal *Candida* overgrowth alone did not increase serum BG. Hence, the detection of BG in serum implied gut-permeability defect in these patients. In addition, BG also induced pro-inflammatory responses through Dectin-1 signaling [12].

With NGS, a high sensitive technology, we could detect bacteria which were unable to grow using standard culture methods. From previous reports, bacterial DNA and RNA were discovered between 4% to 100% in blood of healthy individual [27-31]. Our data agree with Gosiewski et al. which showed increased abundance of proteobacteria in sepsis patients compared to the healthy population (60.1% vs 16.4%) [32]. This raises the possibility that COVID-19 could cause sepsis like syndrome with the same dominant bacteria phylum as in sepsis patients. This phylum contains many genera of bacteria. The most predominant bacteria were *Sphingomonas*, *Bradyrhizobium,* and *Enhydrobacter*. *Sphingomonas paucimobilis* is an opportunistic pathogen that can cause hospital associated infections from environmental exposure [33]. *Bradyrhizobium enterica* were found in patients with colitis [34]. *Enhydrobacter aerosaccus* can be detected from a patient with Hemophagocytic lymphohistiocytosis (HLH) with concomitant corticosteroid use [35]. This is particularly of interest due to the growing evidence of HLH syndrome in severe COVID-19 patients [36].

Patients with COVID-19 have been found to have high levels of proinflammatory cytokines such as IL6, IL-1β, IP10, and MCP-1 [4]. Similar findings were also reported in patients with SARS [37] and MERS-CoV [38]. This has prompted several authors to discuss so-called “*Cytokine Storm*” in viral respiratory infection that is a cause of multiple organ failure. However, our results suggest that bacterial products might be another possible contributor to the cytokine storm rather than only the virus itself.

The source of bacterial toxin, and bacterial DNA in the blood of patients with COVID-19 pneumonia is unclear. It is possible that viremia results in capillary leakage syndrome similar to bacterial sepsis which causes interstitial edema and induces dysfunction of the lung and intestinal barrier which may facilitate bacterial toxin and live bacteria translocation into the circulation. These bacterial products then induce the release of proinflammatory cytokines. In our study, many patients (47.3%) ultimately developed subsequent bacterial infection within 28 days. Hanada et al. proposed the mechanism of viral-induced susceptibility of secondary bacterial infection involving local and systemic immune response which results in alterations in respiratory and gut microbiomes and impaired pulmonary immune response [39]. However, the GI tract appears to be another target of COVID-19. Similar to respiratory tract, various cells in the GI tract also express ACE2 and TMPRSS2 which are crucial for fusion of viral particles with host cells [40, 41]. Biopsies from severe cases have revealed involvement of COVID-19 throughout the GI tract from esophagus to colon [42]. Thus, we propose that loss of gut barrier function might also contribute to the presence of bacterial toxin and bacterial DNA in the blood of patients with severe COVID-19 (Figure 7).

**Figure 7.**
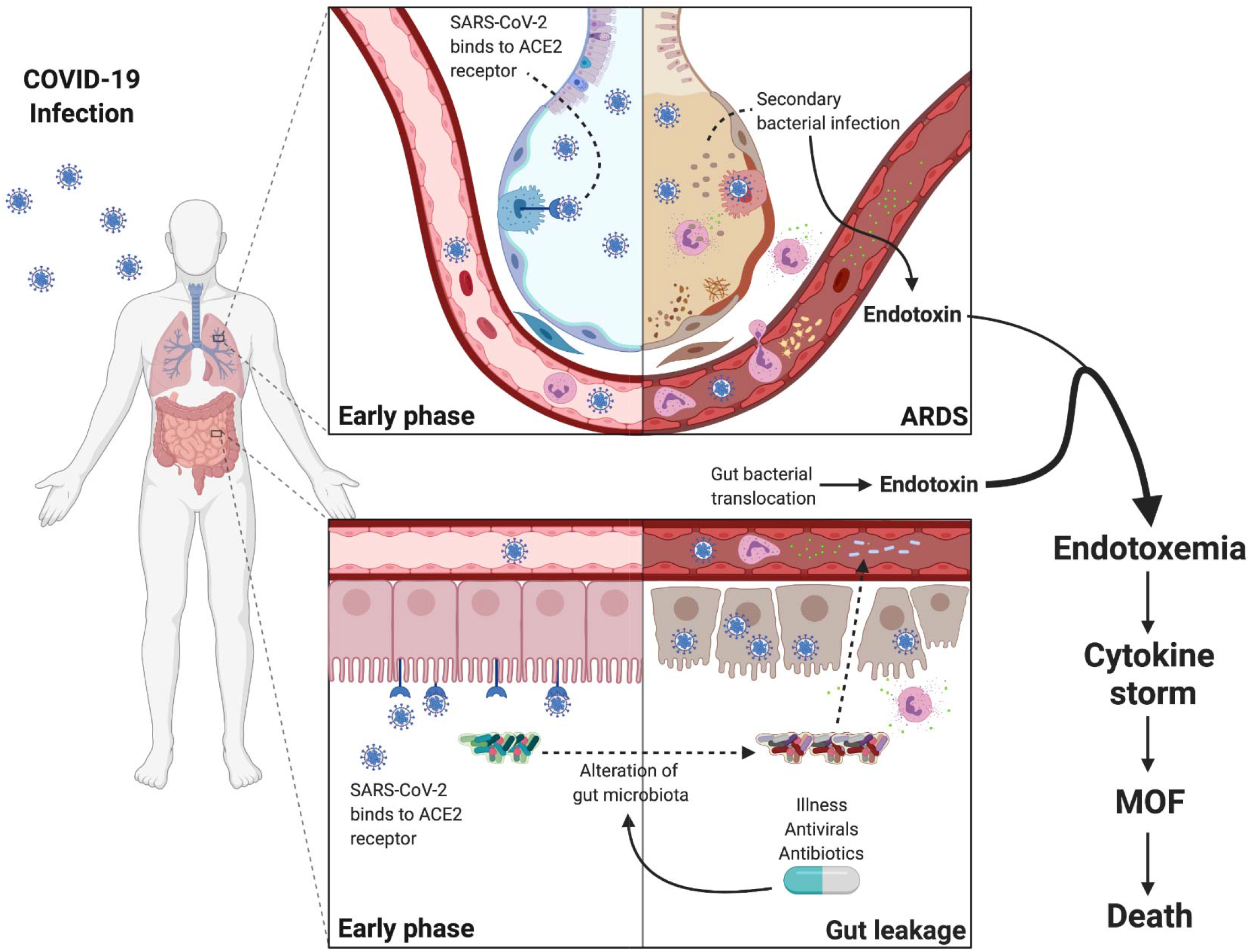
Hypothetical pathogenesis of endotoxemia in COVID-19 pneumonia. At early stage, SARS-CoV-2 primarily infects type 2 pneumocytes in the lungs and causes pneumonia which can progress to ARDS and induces susceptibility of secondary bacterial infection by impairing the pulmonary immune response. The virus can enter the bloodstream causing viremia targeting organs with high ACE2 expression including the gut. SARS-CoV-2 infection of enterocytes causes inflammatory response of gastrointestinal tract which results in alteration of the intestinal microenvironment including epithelial hyperpermeability, attenuated local immune system, and dysbiosis of the microbiome. The perturbations of the intestinal microenvironment allow the pathogenic bacteria from the gut lumen to translocate to the bloodstream. Hence, we propose the sources of endotoxin to be majorly from the gut and minorly from the secondary bacterial infection of the lungs. Abbreviations: COVID-19, the coronavirus disease 2019; SAR-CoV-2, severe acute respiratory syndrome coronavirus 2; ARDS, acute respiratory distress syndrome; ACE2, angiotensin converting enzyme 2; MOF, multiple organ failure

To our knowledge, this is the first study to demonstrate evidence of bacterial toxins including EAA, and the presence of circulating bacteriome in patients with COVID-19 pneumonia. We also showed the association of high level of EAA and the severity of COVID-19 pneumonia. Both the type of bateria and the presence of BG in the serum suggest that the gut is the source.

Our study had several limitations. First, our 16s RNA gene amplification technique could not demonstrate the absolute number of bacterial DNA. Therefore, we could not correlate the burden of circulating bacterial DNA to the severity of the patients. However, our study aimed to be a starting point for future investigation. Second, our study did not include respiratory tract or gastrointestinal specimens, so we cannot completely establish the source of bacterial products in our patients. Future study should explore the effect of organ crosstalk between the lung and the intestine during COVID-19 infection.

## Conclusion

High levels of endotoxin activity and bacterial DNA can be found in the blood of patients with COVID-19 pneumonia which may reflect loss of epithelial barrier function. This previously unrecognized mechanism of hyperinflammation and organ failure in COVID-19 warrants further study.

## Data Availability

The data are not publicly available.

## Contributors

PS, WK, and NS were responsible for study concept and design. PS, WK, NS, TP, AC, SP, PP, SC, VS, NW, SK, OP, TT, NL, VP, NP, TS, RS, ST, KT, NLu, AL, TSri, TTa, UT, CC, KP, KTu, SE, JK, and NS were responsible for the acquisition, analysis, or interpretation of data. PS, WK and NS were responsible for drafting the manuscript. PS, WK and NS were responsible for statistical analysis. All authors had full access to all the data in the study and take responsibility for the integrity of the data and the accuracy of the data analysis. All authors interpreted the findings, contributed to writing the manuscript, and approved the final version for publication.

## Acknowledgments

We would like to thank the staff, fellows, nurses, and research coordinators from the Excellence Center for Critical Care Nephrology (EC-CCN) and Emerging Infectious Disease (EID) unit, Faculty of Medicine, Chulalongkorn University. We also thank Miss Pimnara Peerawaranun for statistical analysis and Miss Sasipha Tachaboon, MSc medical technologist, for laboratory procedures.

---

**S1.**
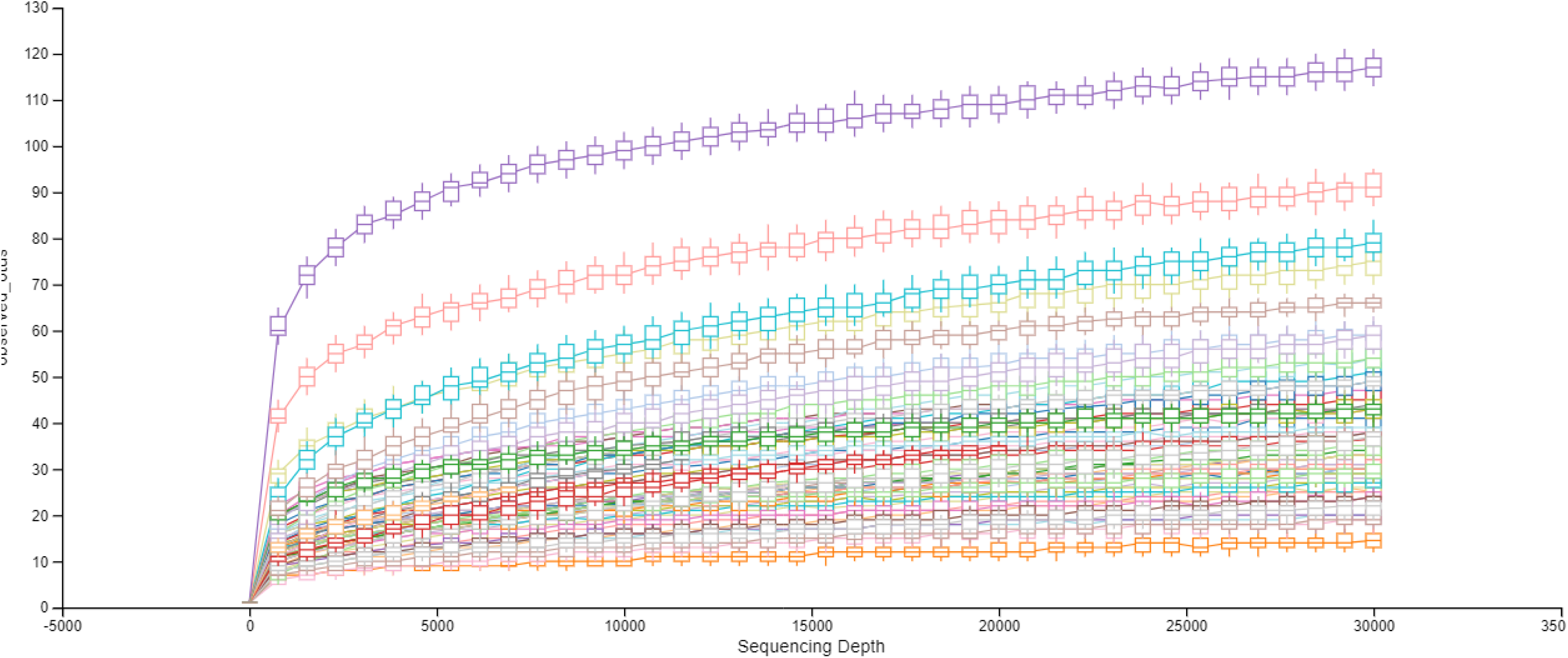
Rarefaction curve of pass-filter reads obobtained from each sample.

**S2.**
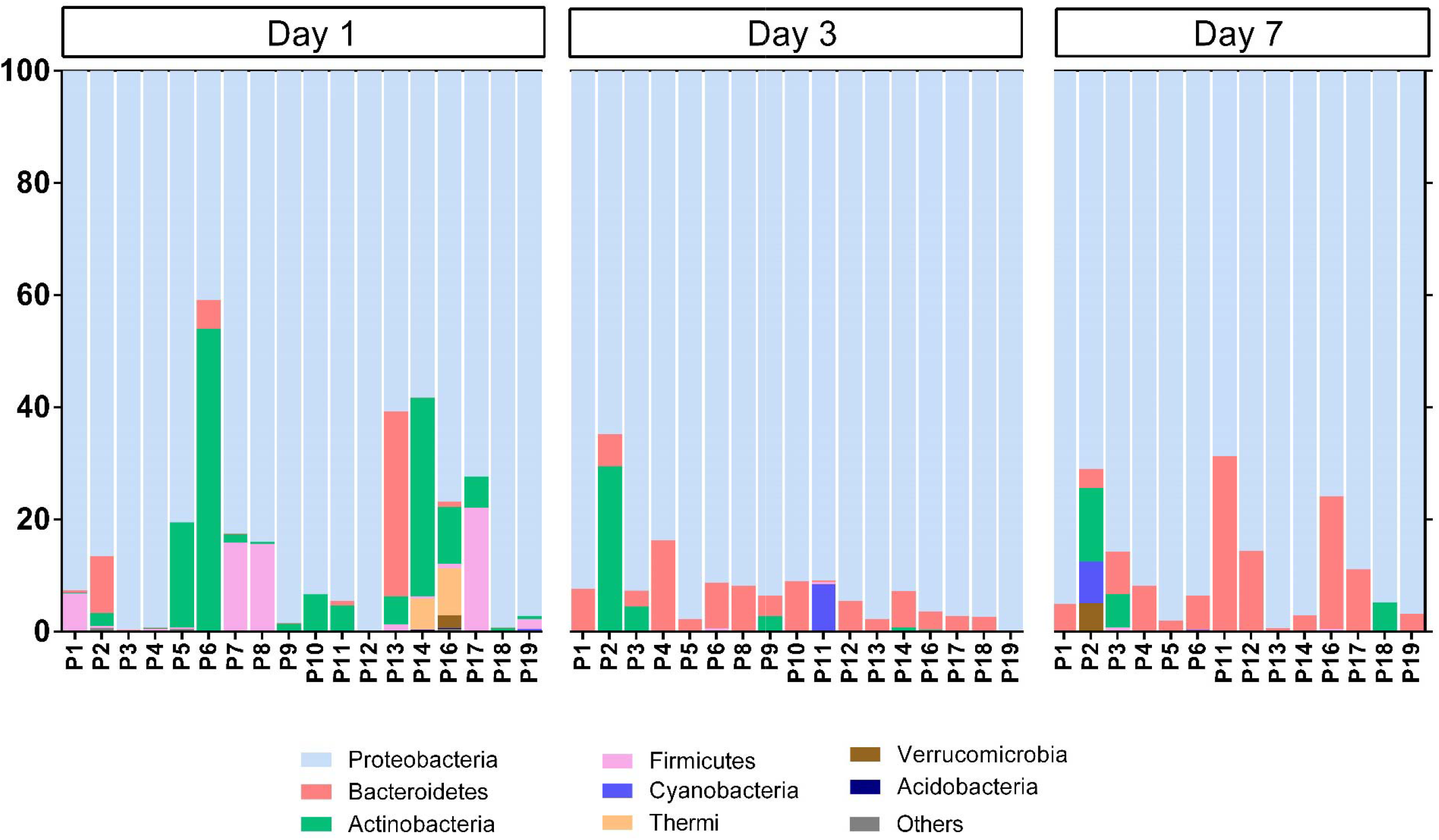
Dynamic bacterial community profiles in in COVID-19 patients on day 1, 3, and 7.

**S3.**
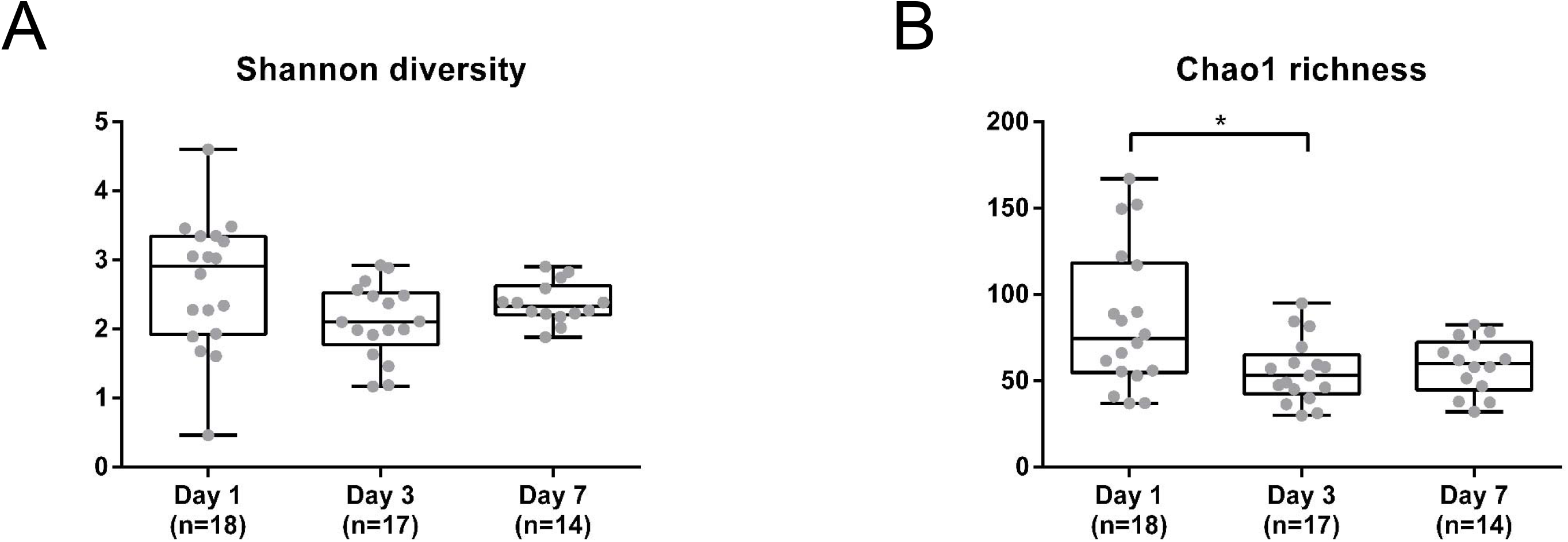
Shannon diversity index (A) and Chao1 richness (B) representing alpha diversity serial community profiles in COVID-19 patientsnts on day 1, 3, and 7.

**S4.**
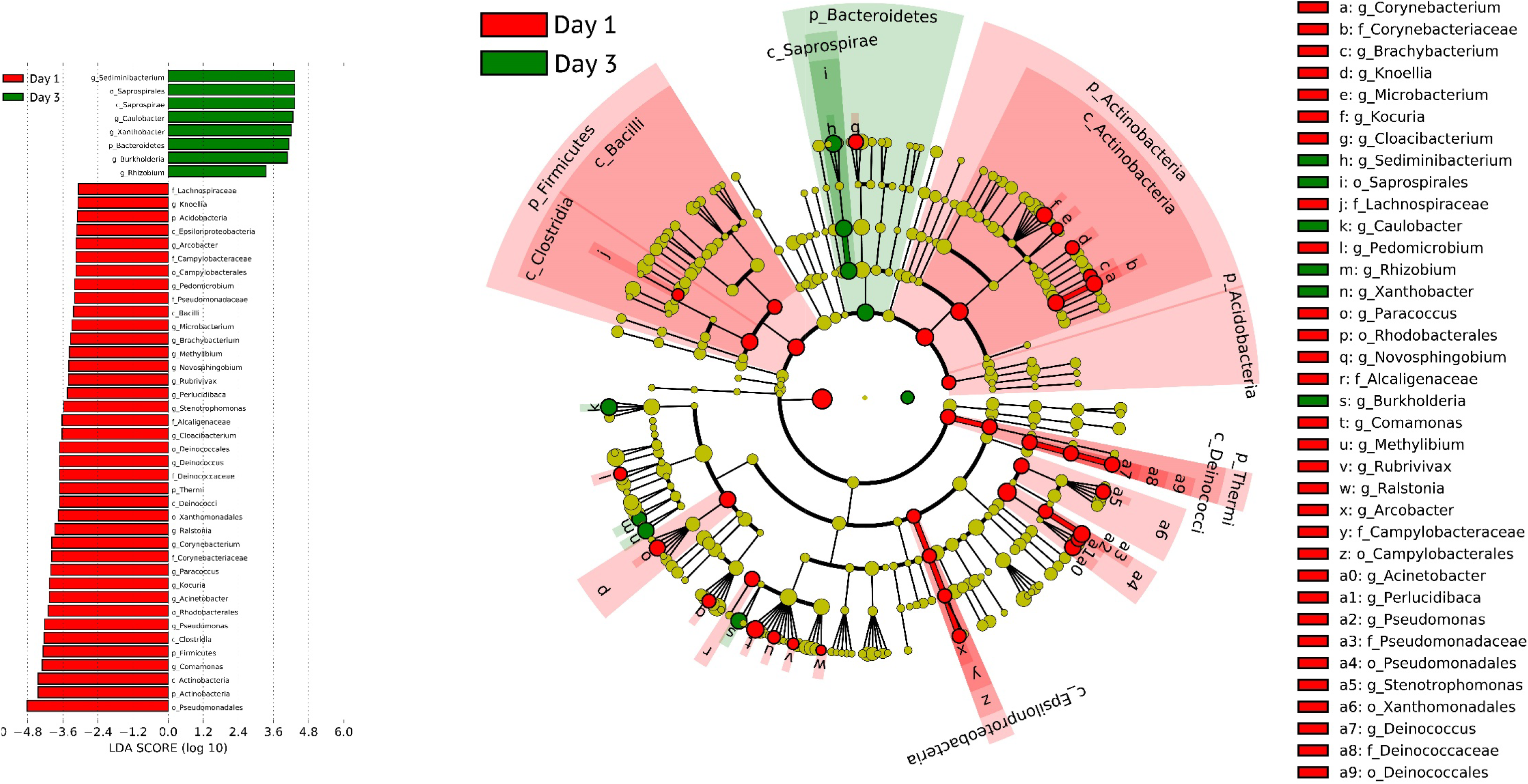
Linear discriminant analysis effect size ((LEfSe) analysis of bacteria on day 1 ay 3.

**S5.**
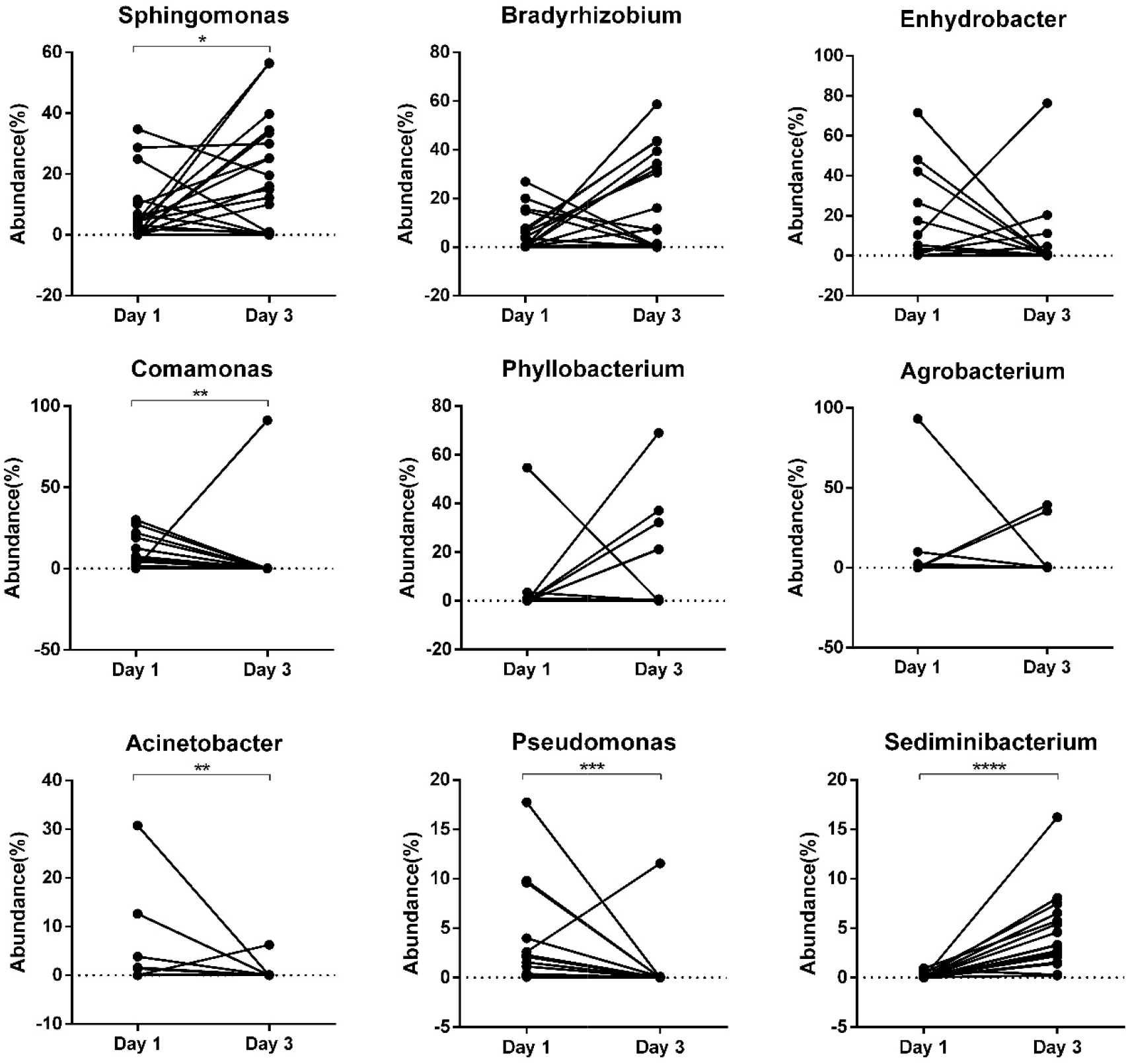
Wilcoxon matched pairs test differentia tial abundant of bacterial genera in each ID-19 patients between day 1 and day 3.

